# Engagement with daily testing instead of self-isolating in contacts of confirmed cases of SARS-CoV-2

**DOI:** 10.1101/2021.03.13.21253500

**Authors:** Alex F Martin, Sarah Denford, Nicola Love, Derren Ready, Isabel Oliver, Richard Amlôt, G. James Rubin, Lucy Yardley

## Abstract

**Background:** In December 2020, Public Health England with NHS Test and Trace initiated a pilot study in which close contacts of people with confirmed COVID-19 were given the option to carryout lateral flow device antigen tests at home, as an alternative to self-isolation for 10-14 days. In this study, we evaluated acceptability of and engagement with daily testing, and assessed levels of adherence to the rules relating to behaviour following positive or negative test results.

**Methods:** We conducted a service evaluation of a pilot study, involving an online cross-sectional survey offered to adult (> 18 years) contacts of confirmed COVID-19 cases who were invited to participate in seven days of daily testing instead of isolation. We used a comparison group of contacts who were not offered testing and performed self-isolation. Herein, we examine survey responses from a subset of those who took part in the pilot study and who responded to the evaluation questionnaire.

**Results:** Acceptability of daily testing was lower among survey respondents who were not offered the option of having it and among people from ethnic minority groups. Overall, 52% of respondents reported being more likely to share details of people that they had been in contact with following a positive test result, if they knew that their contacts would be offered the option of daily testing. Only 2% reported that they would be less likely to provide details of their contacts. On the days that they were trying to self-isolate, 19% of participants reported that they left the house, with no significant demographic group differences. Following a negative test, 13% of respondents reported that they increased their contacts, but most (58%) reported having fewer risky contacts.

**Conclusions:** Our data suggest that daily testing is potentially acceptable, and may facilitate sharing contact details of close contacts among those who test positive for COVID-19, and promote adherence to self-isolation. A better understanding is needed of how to make this option more acceptable for all households. The impact of receiving a negative test on behaviour remains a risk that needs to be monitored and mitigated by appropriate messaging. Future research should examine attitudes and behaviour in a context where infection levels are lower, testing is more familiar, much of the population has been vaccinated and restrictions on activity have been reduced.

## Background

Within the UK, efforts to reduce the spread of SARS-CoV-2 have focussed on the isolation of people who display symptoms of COVID-19, those who test positive for SARS-CoV-2, and their close contacts. This presents a substantial burden for the people involved, financially, socially and psychologically [1]. The isolating of close contacts, most of whom will not be infectious, also generates a wider impact on society and the economy, with sectors deprived of their employees, students, congregants and volunteers. To give an example of the scale involved, in a single week in February 2021 the UK’s NHS Test and Trace service identified 191,242 people (including some duplicates) as having had close contact with someone infected with SARS-CoV-2 [2]. These people were required, by law, to self-isolate for 10 days, meaning that they could not leave their home except for a limited number of reasons (e.g., for an essential medical appointment). Concern exists that even this number is too low, and that many people with confirmed COVID-19 infection do not provide NHS Test and Trace with details of all their contacts [3] perhaps out of concern about the burden self-isolation would place on them.

The extent to which people adhere to the rules relating to self-isolation is unclear. Quantitative estimates provided to date focus on “full adherence.” These studies include people who commit a single minor transgression of the rules which may pose a negligible risk to public health in the same category as people who disregard the rules entirely [4-8]. Despite this limitation, some patterns have emerged from the literature regarding risk factors for non-adherence to self-isolation. These include low income; inability to work from home; and being in jobs that do not provide pay for time off [4]. Periods of lockdown may be particularly challenging for those from lowest income backgrounds (which is over represented by people from minority ethnic groups), who are less able work from home, more likely to be in work where furloughing is not offered, and more likely to experience financial concerns and anxieties than those from less deprived groups [9, 10]. Understanding and supporting adherence to mitigation measures among these populations is critical.

One alternative to self-isolating that has recently been suggested is to ask contacts of cases to undertake a daily coronavirus test using a lateral flow device. If the test result is negative, the individual can continue with their daily activities for a 24 hour period, including leaving their home, provided that they adhere to the governmental policy restrictions that apply for their local area [11]. If the result is positive, the individual must enter 10 days of isolation immediately. If effective and used correctly, this system would dramatically reduce the overall number of days that people spend in self-isolation across the population and may improve case detection. Testing of contacts could be particularly important for maintaining confidence and adherence to the test, trace and isolate system as infection levels decline, since the proportion of false positives in index cases is likely to increase and there will be public awareness that they may be asked to self-isolate when their contact was not actually infectious. However, there are several uncertainties about this proposed system that need to be clarified before it can be used with confidence. Many of these relate to how people use lateral flow devices and whether they adhere to guidance on how to respond to a positive or negative result. Specific challenges include whether people: adhere to guidance to self-isolate while waiting for their test kit to arrive in the post; find the tests easy to use; reduce their adherence to local guidance in the event of a negative result (for example, as a result of false reassurance); and adhere to guidance to isolate in the event of a positive result [12-16].

In December 2020, Public Health England with NHS Test and Trace initiated a pilot study in which close contacts of confirmed cases of COVID-19 were given the option to either enter a 10-14 day self-isolation period as normal or to undertake consecutive lateral flow device antigen tests during the first 7 days post exposure. A report on the testing element of the study has been provided elsewhere [17]. For this study, we carried out a survey to evaluate the acceptability of daily testing and assess levels of adherence to the rules relating to behaviour following a positive and negative test result in a subset of individuals who performed serial testing. Our specific objectives were to:

- Investigate whether the offer of daily testing would motivate people who have COVID-19 to provide details about more contacts to NHS Test and Trace;
- Understand the reasons for accepting daily testing, any issues people have conducting daily testing, and levels of confidence in daily testing;
- Assess levels of adherence to the rules, including leaving the house and number of non-household contacts, following positive or negative test results;
- Determine the proportion of contacts who reported close contact with other people or activities outside the home when they had a positive, negative or inconclusive test and when they were trying to self-isolate.

## Method

This study examines data which was part of a larger study where asymptomatic contacts of cases (recruited between 11 and 23 December 2020 and 4 to 12 January 2021) received six lateral flow devices (LFD), a PCR self-sample postal swab kit and study documentation.

They were asked to complete up to six tests and report results daily to a PHE data collection portal. The findings of this study are reported elsewhere [17].

## Design

We conducted an ethically approved service evaluation (granted by Public Health England Research Ethics and Governance Group: Reference NR0235), involving an online cross-sectional survey of adult contacts of confirmed COVID-19 cases who were invited to participate in seven days, post exposure daily testing, as an alternative to 10-14 days isolation.

Participants who agreed to daily testing were posted a study testing pack consisting of 6 lateral flow devices, a PCR self-sample postal swab kit and study documentation. A text message was sent to all participants with a valid mobile number to inform them that their kit had been sent, and to provide participants with the hyperlink to a “results portal” where they were required to submit daily test results, a digital image of the test, identifiers (name, date of birth, postcode, NHS number), a record of any symptoms experienced and the date the kit arrived. Participants were asked to complete up to 6 tests and report results to PHE each day using the results portal.

### Survey recruitment

A survey was sent via text message to all consenting participants recruited into the main study if they had a valid mobile phone number. Individuals were divided into three groups: those offered daily testing and who accepted it, those offered daily testing but who declined it, and those eligible but not offered daily testing.

Figure one shows the flow of participants through the study. Of the 1760 individuals offered daily testing, 882 agreed to it and 878 declined. A total of 923 individuals consented to further contact from NHS Test and Trace for the purposes of our service evaluation. As a non-randomised comparison group, 857 individuals who were eligible for inclusion in the daily testing trial but who were not offered it for capacity reasons, and who had agreed to further contact from NHS T&T, were also sent a link to the evaluation questionnaire. This group was matched to those offered daily testing by age and the date daily testing was offered.

A total of 668 surveys were returned. Although the survey could be completed anonymously, participants had the option to provide identifiable information (names and NHS Test and Trace identifying number). Where participants did not provide these details (N = 332), we checked if they had answered the correct sections of the questionnaire (checking, for example, if participants offered testing had completed items suggesting they had taken tests). We could not confirm group allocation for 74 participants, and these participants were excluded. A total of 21 participants completed the survey on multiple occasions (e.g., after reminder emails were sent to all respondents). For these participants, we retained only their first set of answers for the analyses. We also excluded 72 participants who reported that they did not receive a testing kit in time to participate.

Only 12 participants out of 142 who provided consent (1.4%) provided usable data in the “declined testing” group. We therefore excluded this group from all analyses. Usable data were included from 319 people who had agreed to daily testing (out of 781 who provided consent, 36.2%) and 205 who were not offered daily testing (24.0%).

### Study materials

All those consenting to further contact were sent a link to an electronic evaluation questionnaire, developed in Snap Survey. Survey questions were filtered depending on participants’ self-selected group allocation (accepted, declined or not offered daily testing). Full survey materials are available in Appendix 1.

#### Demographics

We asked participants to report their age, gender, highest educational qualification and ethnicity.

#### Preference for daily testing and sharing contacts

We asked participants to rate their preference for daily testing on a five-point item (strongly prefer testing to strongly prefer self-isolating). We asked them to rate how likely they would be to share contact details of close contacts if the option of daily testing (instead of self-isolating) was available to their contacts (much more likely to much less likely).

#### Perceptions of daily testing

We asked participants who had accepted daily testing to select from prescribed options their reasons for daily testing and any problems that made it hard to complete the tests and submit the results, more than one option could be selected. Participants were also asked if they had to repeat tests (yes or no).

We asked these participants to rate on five-point items how confident they were that they had taken the tests correctly, and how confident they were in the test results (very confident to not at all confident).

#### Activities and contacts

Participants who completed daily testing were asked to report a test result (positive, negative or invalid) on each day that they took a test. They were also asked to report the number of times that they came into close contact with someone that they did not live with (0 times, 1 time, 2-4 times, 5-10 times, and 11 or more times) on each day during the seven day, post exposure testing period.

We asked participants to state whether or not they had left their home 1) on days that they were trying to self-isolate (which should have included days spent waiting for the test kit to arrive and any days after testing positive) and 2) on days that they had a negative test result. For both of these, participants could select from: 1) to go to the shops for groceries, toiletries or medicine; 2) to go to the shops for other items; 3) to go to work, school or university; 4) to help or provide care for someone; 5) to spend time indoors and in close contact (less than a meter apart and for more than 15 minutes) with friends or family that they did not live with; 6) to go out for a meal or to an entertainment venue; 7) to take a child to or from school; 8) to exercise; 9) to attend a medical appointment; 10) or for any other reason. Participants could also select 11) ‘did not go out for any reason’ as an option.

Finally, we asked participants to rate on a five point item the frequency of close contacts (with people you do not live with, indoors and for more than 15 minutes; much more contact to much less contact) on the days when they were trying to self-isolate, and on the days when they had a negative test result.

#### Ethics

Ethical approval for this study was granted by Public Health England’s Research Ethics and Governance Group (Reference NR0235).

### Data analysis

We split the sample into those who accepted testing and self-reported that they received at least one LFD positive result from a lateral flow device (PosTest N = 54), those who accepted LFD testing and did not self-report a positive result (NegTest N = 265), and those who were not offered daily LFD testing (Not Offered N = 205). As we had low group sizes and because we were interested in examining those with lower levels of education and those from an ethnic minority background, we created two binary groups. Education was collapsed into secondary education (did not complete school or finished education after School; N = 289) and higher education (completed university or postgraduate degree; N = 163). Ethnicity was collapsed into ethnic minority (N = 42) and White (N = 440). Age was collapsed into five categories (18-24; 25-34; 35-44; 45-54; 55+ years).

To explore acceptability of daily testing, we present proportions of respondents selecting each option for items on their preference for daily testing or self-isolating and the likelihood of providing details of contacts item by testing groups and by demographic group.

To explore behaviour, we used three measures. First, we coded behavioural activities as higher risk non-essential contacts (items 1-6, 10), lower risk non-essential contacts (items 7-8), or no non-essential contacts (items 9 and 11) and report proportions when self-isolating and following a negative test result. Second, we report whether people reported more or less close contact when self-isolating or following a negative test result. Third, we present the frequency of contacts following a positive test result.

Finally, we present proportions of respondents selecting each option of the two confidence items and proportions of participants selecting each option regarding motives for accepting daily testing.

Chi square tests were used to examine differences in proportions between the groups. Data were analysed using R version 3.4.3 [18]

## Results

### Demographics

Demographic breakdowns are presented in Table 1. In the Not Offered group there was a higher proportion of participants self-reporting to be in the higher education category and a higher proportion of women than in the PosTest and NegTest groups. In the PosTest group, data suggested that there was a higher proportion of participants from ethnic minority groups than in the NegTest or Not Offered groups, and a higher proportion of participants in the secondary education category.

**Table 1.** Demographic characteristics of the whole sample and by group

|  | Whole<br>sample | % | Pos<br>Test | % | Neg<br>Test | % | Not<br>offered | % |
| --- | --- | --- | --- | --- | --- | --- | --- | --- |
| Group N | 524 | - | 54 | 10% | 265 | 51% | 205 | 39% |
| Ethnicity |  |  |  |  |  |  |  |  |
| BAME | 42 | 8% | 8 | 15% | 18 | 7% | 16 | 8% |
| White | 440 | 84% | 40 | 74% | 229 | 86% | 171 | 83% |
| No response | 42 | 8% | 6 | 11% | 18 | 7% | 18 | 9% |
| Sex |  |  |  |  |  |  |  |  |
| Female | 266 | 51% | 21 | 39% | 116 | 44% | 129 | 63% |
| Male | 170 | 32% | 16 | 30% | 90 | 34% | 64 | 31% |
| No response | 88 | 17% | 17 | 31% | 59 | 22% | 12 | 6% |
| Education |  |  |  |  |  |  |  |  |
| Secondary<br>education | 289 | 55% | 28 | 52% | 85 | 32% | 62 | 30% |
| Higher education | 163 | 31% | 16 | 30% | 145 | 55% | 116 | 57% |
| No response | 72 | 14% | 10 | 19% | 35 | 13% | 27 | 13% |
| Age |  |  |  |  |  |  |  |  |
| 18-24 years | 33 | 6% | 0 | 0% | 20 | 8% | 13 | 6% |
| 25-34 years | 94 | 18% | 7 | 13% | 51 | 19% | 36 | 18% |
| 35-44 years | 107 | 20% | 16 | 30% | 59 | 22% | 32 | 16% |
| 45-54 years | 126 | 24% | 16 | 30% | 68 | 26% | 42 | 20% |
| 55+ years | 146 | 28% | 14 | 26% | 55 | 21% | 77 | 38% |
| No response | 18 | 3% | 1 | 2% | 12 | 5% | 5 | 2% |

### Preference for daily testing and sharing contacts

Of the 1760 participants offered daily testing, 882 accepted. Of the 878 who declined, 343 (39.1%) reported that this was because they already had access to testing. Adjusting for this, of the 1,417 people who had not accessed testing, 882 (62.2%) accepted daily testing. Participants’ preferences for daily testing or self-isolation are presented in Figures 2 and 3. Individuals who eligible but were not offered daily testing appeared most divided of the groups, with 46% preferring or strongly preferring testing and 41% preferring or strongly preferring isolation. Participants from ethnic minority communities were similarly divided, with 48% preferring or strongly preferring testing and 48% preferring or strongly preferring isolation (compared with participants identifying as White, with 70% preferring or strongly preferring testing and 23% preferring or strongly preferring isolation).

**Figure 1.**
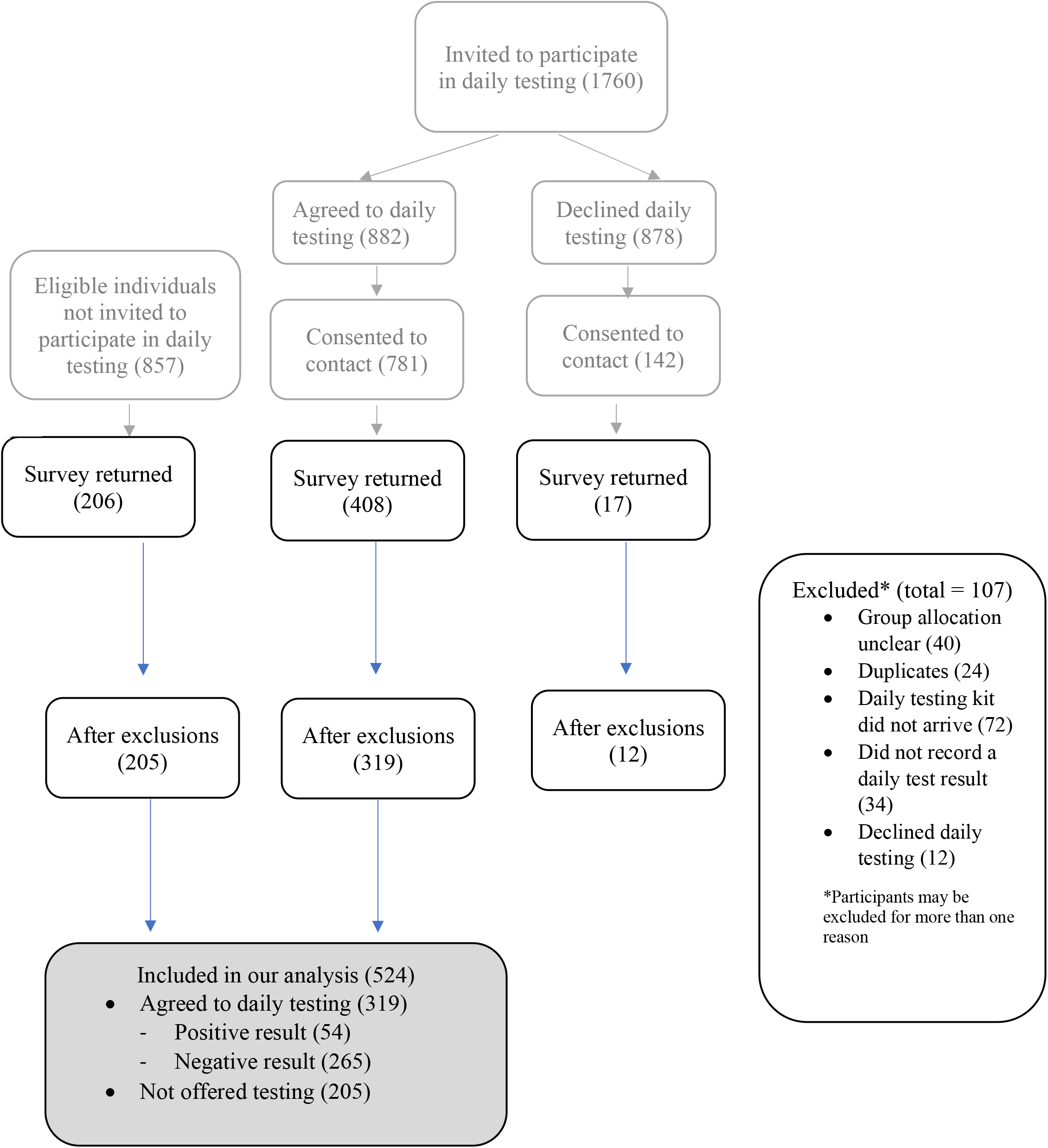
Flow chart showing participants invited to take part, consenting to take part in the study, responding to surveys, exclusions, and final group count

**Figure 2.**
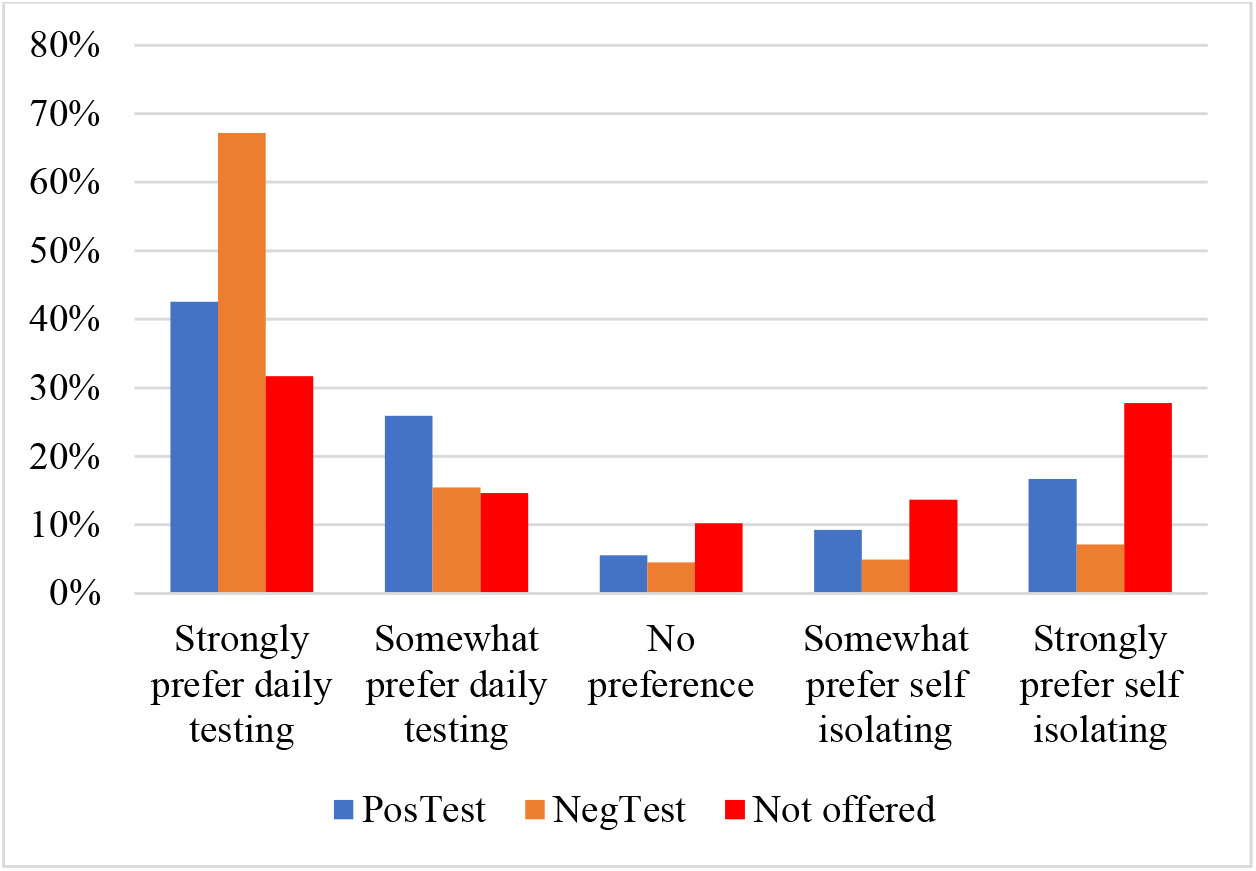
Preference for daily testing or self isolating, by isolation group

**Figure 3.**
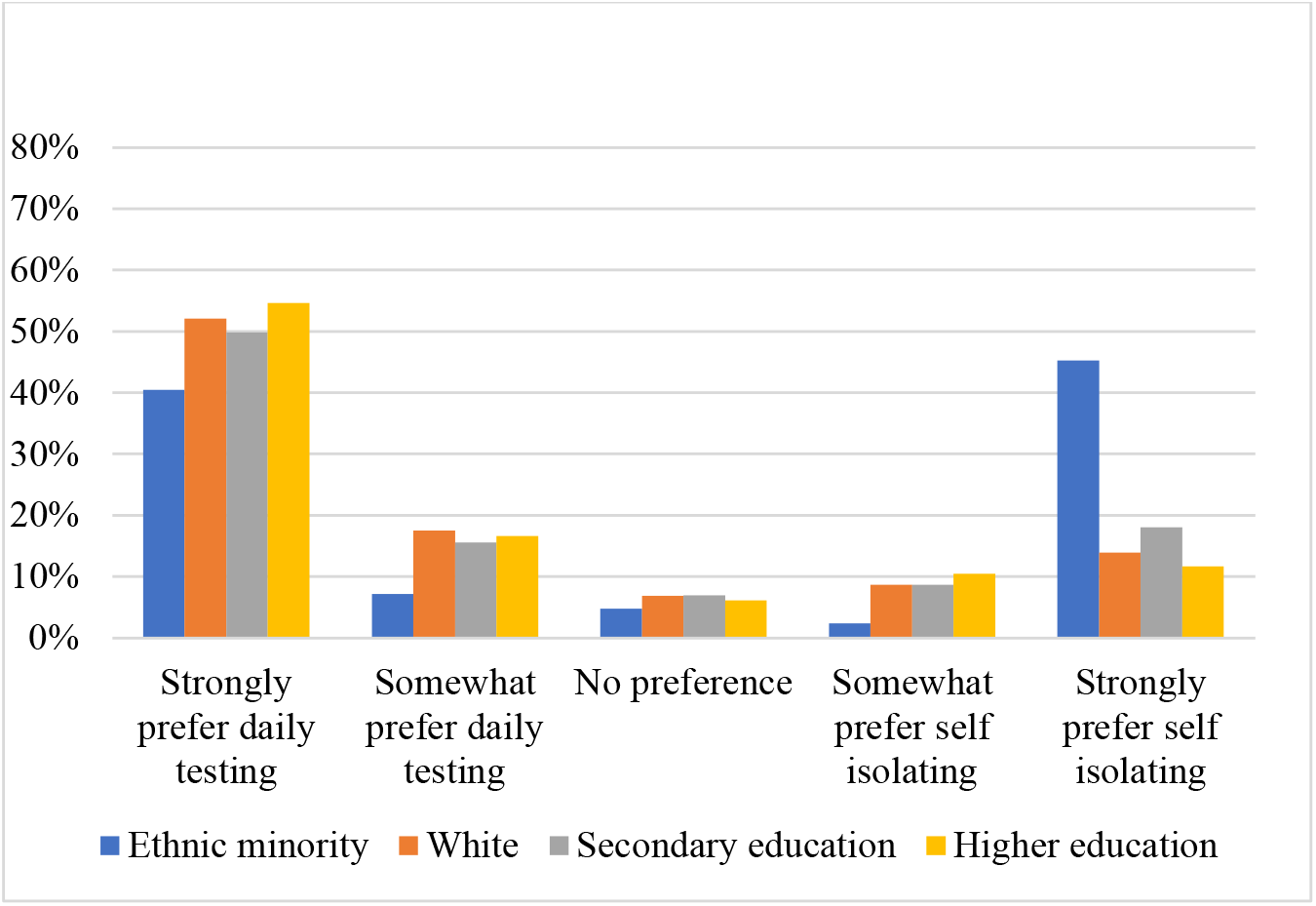
Preference for daily testing or self isolating, by demographic group

Participants also reported that the availability of daily testing would encourage sharing of contact details, with 52% of the sample reporting that they would be more likely to share the details of people that had been in contact with following a positive test result, and 44% reporting that it would make no difference (Table 2). Only nine people across the whole sample (2%) reported that it would make them less like to share contact details.

**Table 2.** Likelihood of sharing contacts if daily testing was available

|  | Whole<br>sample | % | Pos<br>Test | % | Neg<br>Test | % | Not<br>offered | % |
| --- | --- | --- | --- | --- | --- | --- | --- | --- |
| Much more likely | 224 | 43% | 22 | 41% | 135 | 51% | 67 | 33% |
| Somewhat more likely | 49 | 9% | 9 | 17% | 24 | 9% | 16 | 8% |
| It would make no<br>difference | 228 | 44% | 21 | 39% | 100 | 38% | 107 | 52% |
| Somewhat less likely | 4 | 1% | 0 | 0% | 1 | 0% | 3 | 1% |
| Much less likely | 5 | 1% | 0 | 0% | 1 | 0% | 4 | 2% |
| Don't know | 14 | 3% | 2 | 4% | 4 | 2% | 8 | 4% |

\* Item: "If you had a positive test in the future and you knew that your contacts would be able to have daily testing (instead of self-isolating), would you be more or less likely to give their contact details?"

### Perceptions of daily testing

The most commonly given reasons for accepting daily testing as reported in the survey were: I wanted to know if I had the virus (22%); To help beat the virus in my area (20%); It sounded easy to do (17%); and I needed to know if I was infected so I could protect vulnerable people that I live with or meet regularly (13%). This was broadly similar between ethnicity groups and education categories, however, participants from ethnic minority communities also reported: I was concerned that I might have coronavirus (11%).

Two thirds (67%) of participants reported that they had no problems with daily testing. Of those who did report any issues (N = 108, 20%), the most frequent responses were: Internet/technology access (6%); The testing procedure was unpleasant (4%); and Instructions were not clear (4%). Only 2% of those in the daily testing groups had to repeat tests due to inconclusive results.

Of the participants in the daily testing groups, 88% were completely or very confident that they did the test correctly, and 68% were completely or very confident that the test was accurate (Table 3).

**Table 3.** Confidence in tests (daily testing groups)

|  | Did test<br>correctly | % | Test was<br>accurate | % |
| --- | --- | --- | --- | --- |
| Completely<br>confident | 163 | 51% | 101 | 32% |
| Very confident | 119 | 37% | 117 | 37% |
| Fairly confident | 32 | 10% | 79 | 25% |
| Not very confident | 5 | 2% | 5 | 2% |
| Not at all confident | 0 | 0% | 14 | 4% |
| Don't know | 0 | 0% | 4 | 1% |

Item: "How confident, if at all, are you that your test results were accurate?"

### Activities and contacts

Most participants reported not engaging in any non-essential activities on days when they were trying to isolate (80% PosTest, 82% NegTest and 83% Not Offered; Table 4). There were no observed differences between the groups (*𝒳* = 1.64, *p* = .199).

**Table 4.**
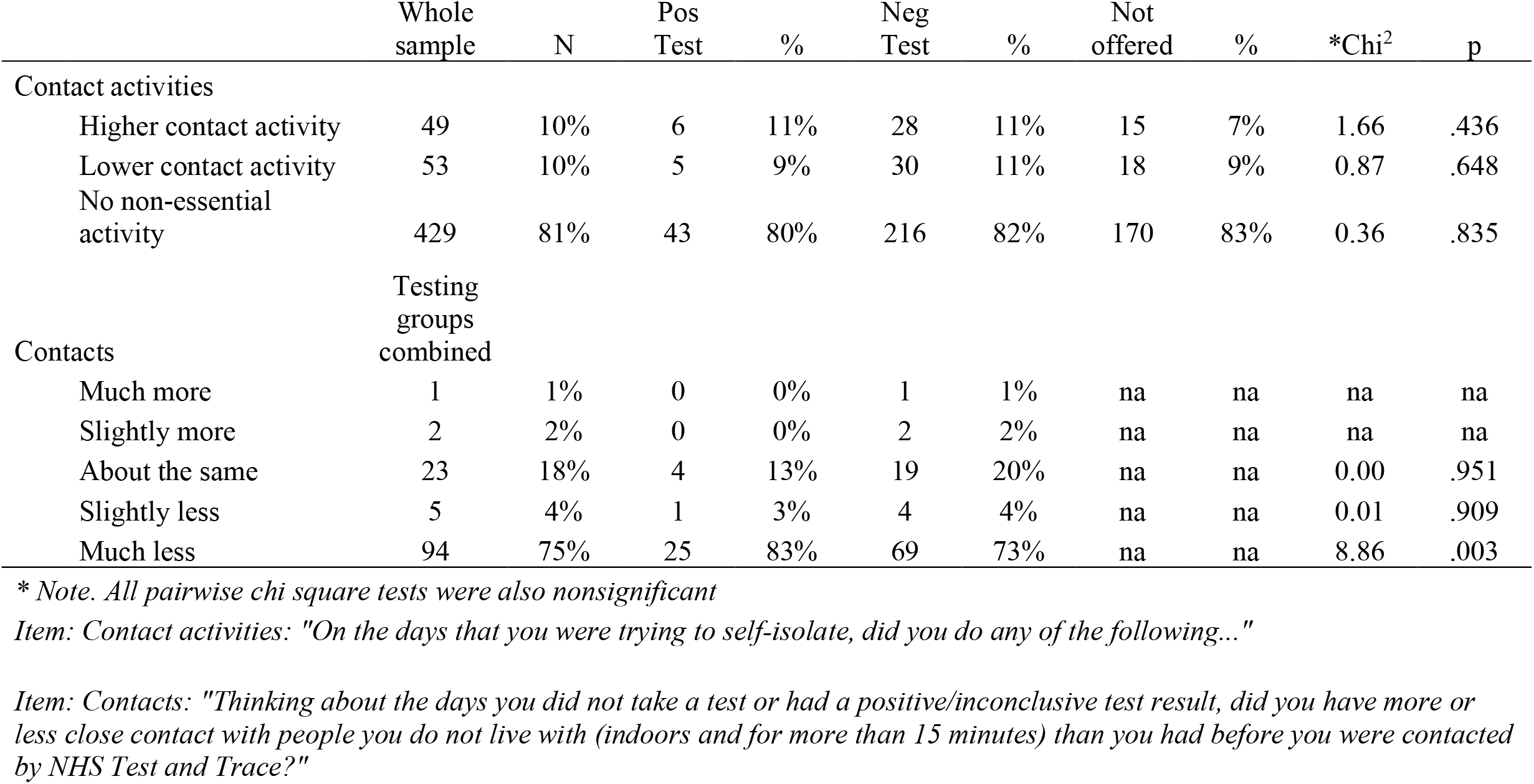
Contact activities when self isolating, by group

|  | Whole<br>sample | N | Pos<br>Test | % | Neg<br>Test | % | Not<br>offered | % | *Chi<sup>2</sup> | p |
| --- | --- | --- | --- | --- | --- | --- | --- | --- | --- | --- |
| Contact activities |  |  |  |  |  |  |  |  |  |  |
| Higher contact activity | 49 | 10% | 6 | 11% | 28 | 11% | 15 | 7% | 1.66 | .436 |
| Lower contact activity | 53 | 10% | 5 | 9% | 30 | 11% | 18 | 9% | 0.87 | .648 |
| No non-essential<br>activity | 429 | 81% | 43 | 80% | 216 | 82% | 170 | 83% | 0.36 | .835 |
| Contacts | Testing<br>groups<br>combined |  |  |  |  |  |  |  |  |  |
| Much more | 1 | 1% | 0 | 0% | 1 | 1% | na | na | na | na |
| Slightly more | 2 | 2% | 0 | 0% | 2 | 2% | na | na | na | na |
| About the same | 23 | 18% | 4 | 13% | 19 | 20% | na | na | 0.00 | .951 |
| Slightly less | 5 | 4% | 1 | 3% | 4 | 4% | na | na | 0.01 | .909 |
| Much less | 94 | 75% | 25 | 83% | 69 | 73% | na | na | 8.86 | .003 |

\* Note. All pairwise chi square tests were also nonsignificant
Item: Contact activities: "On the days that you were trying to self-isolate, did you do any of the following..."
Item: Contacts: "Thinking about the days you did not take a test or had a positive/inconclusive test result, did you have more or less close contact with people you do not live with (indoors and for more than 15 minutes) than you had before you were contacted by NHS Test and Trace?"

As expected, participants engaged in more non-essential activities following a negative test result than on the days that they were trying to self-isolate (Table 5). Participants in the PosTest group appeared less likely to engage in non-essential activities following a negative test result than those in the NegTest group. They also reported fewer lower contact activities following a negative test result than those in the NegTest group (15% versus 49%, *𝒳* = 21.89, *p* = < .001).

**Table 5.** Contact activities with a negative test result, by group

|  | Whole<br>sample | N | Pos<br>Group | % | Neg<br>Group | % | Not<br>offered | % | *Chi<sup>2</sup> | p |
| --- | --- | --- | --- | --- | --- | --- | --- | --- | --- | --- |
| Contact activities |  |  |  |  |  |  |  |  |  |  |
| Higher contact activity | 88 | 26% | 10 | 19% | 78 | 29% | na | na | 2.68 | .102 |
| Lower contact activity | 139 | 41% | 8 | 15% | 131 | 49% | na | na | 21.89 | < .001 |
| No non-essential<br>activity | 111 | 33% | 30 | 56% | 81 | 31% | na | na | 12.35 | < .001 |
| Contacts | Testing<br>groups<br>combined |  |  |  |  |  |  |  |  |  |
| Much more | 7 | 3% | 0 | 0% | 7 | 4% | na | na | na | na |
| Slightly more | 15 | 7% | 1 | 4% | 14 | 7% | na | na | 1.18 | .278 |
| About the same | 63 | 30% | 6 | 24% | 57 | 30% | na | na | 3.06 | .080 |
| Slightly less | 20 | 9% | 1 | 4% | 19 | 10% | na | na | 2.19 | .142 |
| Much less | 108 | 51% | 17 | 68% | 91 | 48% | na | na | 0.16 | .686 |

\* Note. All pairwise chi square tests were also nonsignificant
Item: Contact activities: "On the days that you had a negative test result, did you do any of the following ..."
Item: Contacts: "Thinking about the days you had a negative result, did you have more or less close contact with people you do not live with (indoors and for more than 15 minutes) than you had before you were contacted by NHS Test and Trace?"

Among those who tested negative, only 20 people (13%) reported engaging in more high risk activity (i.e. indoor close contact for more than 15 minutes) than prior to testing, and most (58%) reported having fewer risky contacts than they had before they were contacted by NHS Test and Trace.

Out of the 54 people who reported a positive test, 7 (13%) reported having close contact with people that they did not live with following the positive test. Of these seven, two reported contacts with only a single person on one day. The other five reported multiple contacts (the categories were: ‘two to four’, ‘five to ten’ and ‘eleven or more’).

## Discussion

To the best of our knowledge, this is the first study to investigate the acceptability of daily testing of confirmed COVID-19 cases, and to attempt to understand the impact of testing on activities and contacts. Participants from White backgrounds reported a strong preference for daily testing over 10 days isolation. However, participants who were not offered the option of daily testing and those from ethnic minority groups were more divided. These findings suggest that if daily home testing is offered there is a need to develop materials and campaigns to explain the rationale and procedures and address concerns, especially among ethnic minority communities.

In addition to exploring whether or not daily testing is preferable to self-isolation, we also explored whether the option of daily testing would encourage positive cases to share the contact details of their close contacts. Contact tracing is dependent on the willingness of positive cases to share the details of those they have been in contact with, and individuals may be reluctant to cause disruption to their friends and family [19]. Over half the participants who completed this survey reported that knowing that their contacts would have the option of daily testing would make them more likely to provide their details, and most other participants reported that it would make no difference. Very few participants reported that it would make them less likely to share contact details of close contacts. If obtaining the details of more contacts of cases is a priority for NHS Test and Trace, or other agencies around the world, the offer of daily testing may go some way towards achieving this goal. Overall, participants were confident in their ability to perform the tests correctly and two-thirds of participants were very, or completely, confident in the accuracy of the lateral flow test results. At the time this study was carried out there was considerable public debate in the media about the accuracy of lateral flow tests, which may have contributed to concerns. In future studies, further explanation of the benefits and limitations of testing could be provided. The most common motives for taking daily tests were to find out if you had the virus, to help beat the virus, and to protect vulnerable people that they lived with or met regularly. Participants also reported that they wanted to take daily tests because it sounded easy to do. In addition, those from ethnic minority groups reported wanting to take part in daily testing because they were concerned that they may have the virus. Two thirds of participants reported having no issues with daily testing. The most common problems reported by participants included unclear instructions, tests being unpleasant, and IT/internet issues. While the unpleasantness of tests is perhaps unavoidable, further work to clarify the instructions and provide alternative, non-internet-based routes to certify a test result may be beneficial.

Most participants in the survey reported that they either had less close contact, or about the same amount of close contact, with people outside of their household than before they were contacted by NHS test and trace. In contrast to previous research [5] the majority of participants who completed the survey reported that they did not leave the house for any reason on the days that they were trying to self-isolate (either in response to a positive test result, or whilst waiting for testing kits to arrive). This may be in part because participants were recruited for daily testing when infection rates were extremely high, and many participants had their daily activities limited by the restrictions in place in their local areas. Nonetheless, it was notable that those with a positive test result during the seven day testing period reported engaging in fewer contact activities compared to those who only had negative tests. Seven people (13%) reported contact with people they did not live with following a positive test result, sometimes on multiple occasions, although it is not clear what the reasons for this were. In future research, it will be important to examine reasons for non-adherent behaviour following a positive test. The behaviour following a negative test result of participants in our study is consistent with data generated as part of mass testing in Liverpool, where 17% of respondents reported being more likely to go to the shops, and 9% more likely to visit friends and family, following a negative test [16].

This study has several limitations. First, the overall response rate was low. It is possible that those who completed the survey are not representative of the general population, and may have been more adherent than those who declined to take part in the survey. In particular, the response rate of participants who declined to take part in daily testing was very low, and as a result this group had to be excluded from the analyses. Comparisons between those who accepted daily testing and the comparison group who were not offered testing are problematic as the comparison group was non-randomised and only partially matched, and had a lower uptake, with a higher proportion of women and those with higher education levels participating. Second, all data were self-reported and may therefore have been susceptible to social desirability bias. Third, the fact that some participants were invited to take part in daily testing over the Christmas may have had an impact on the study. For example, activity patterns may have been unusual. Finally, daily testing was introduced so that individuals who have a negative test result do not have to isolate. At the time that daily testing was introduced, many areas of the UK were in Tiers three or four, meaning that substantial restrictions were in place. On the 5^th^ January 2021, the government introduced a third national lockdown and all non-essential businesses had to close. It is possible that participants were less willing to take daily tests during lockdown if the need to leave the home was reduced.

## Conclusion

Overall, our data suggested that daily testing has the potential to be an acceptable alternative to self-isolation. However, there is a need to develop materials and campaigns to explain the rationale and procedures and address concerns, especially among BAME communities. Our data also suggests that daily testing may facilitate sharing contact details of close contacts among those who test positive for COVID-19, and could promote adherence to self-isolation. While receiving a negative test result did not appear to lead to substantially increased activity in this study, this remains a risk that needs to be monitored and minimised as far as possible by appropriate messaging. Further research is now needed to explore the uptake and efficacy of daily testing among a wider range of individuals outside of lockdown.

## Data Availability

The datasets used and/or analysed during the current study are available from the corresponding author on reasonable request.

## Declarations

### Ethics approval and consent to participate

Ethical approval for this study was granted by Public Health England Research Ethics and Governance Group (Reference NR0235)

### Consent for publication

All participants provided verbal consent for data to be included in publications

## Authors’ contributions

Conceived the study: All authors

Study design: All authors

Analysed the data: AM

Interpreted the data: All authors

Drafted the manuscript: AM and SD

Reviewed the manuscript and approved content: All authors

Met authorship criteria: All authors

## Acknowledgements

Lucy Yardley is an NIHR Senior Investigator and her research programme is partly supported by NIHR Applied Research Collaboration (ARC)-West, NIHR Health Protection Research Unit (HPRU) in Behavioural Science and Evaluation, and the NIHR Southampton Biomedical Research Centre (BRC).

Sarah Denford is supported by the NIHR Health Protection Research Unit (HPRU) in Behavioural Science and Evaluation at the University of Bristol in partnership with Public Health England.

Alex F Martin is supported by the Economic and Social Research Council Grant Number ES/J500057/1 and the NIHR HPRU in Emergency Preparedness and Response at King’s College London in partnership with Public Health England.

James Rubin is supported by the NIHR HPRU in Emergency Preparedness and Response at King’s College London in partnership with Public Health England.

## Supplementary materials

**Supplementary table 1.**
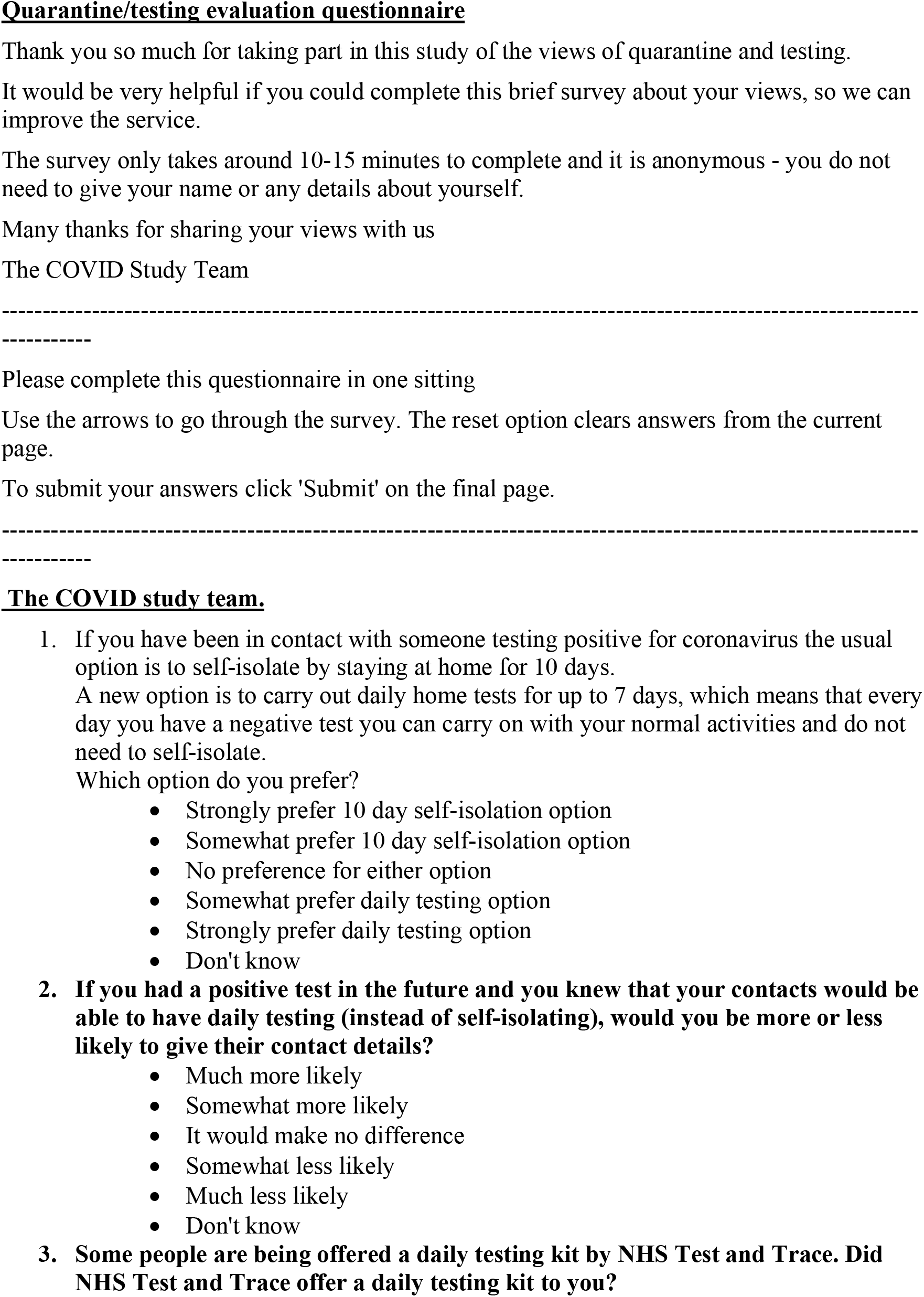
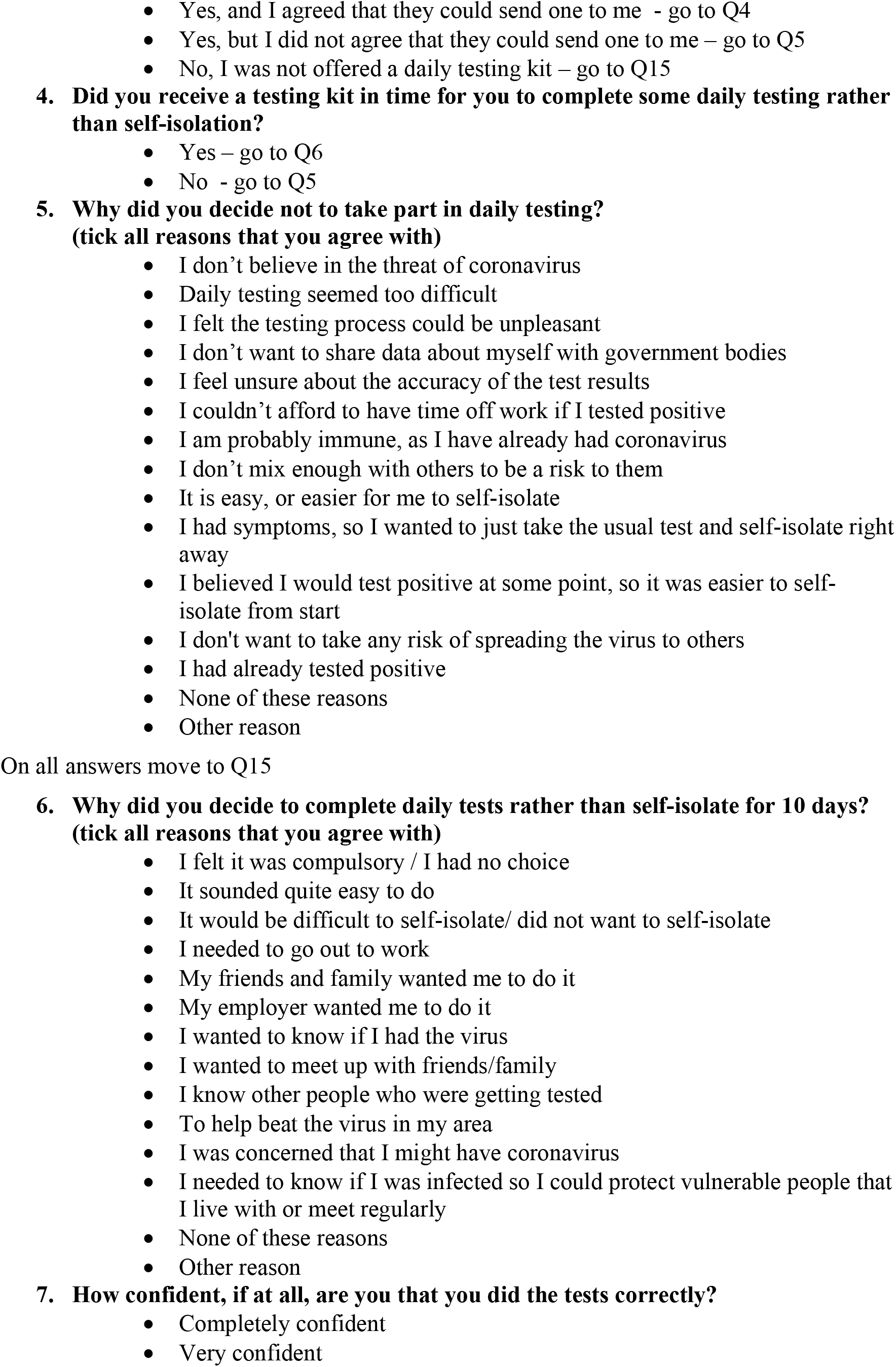
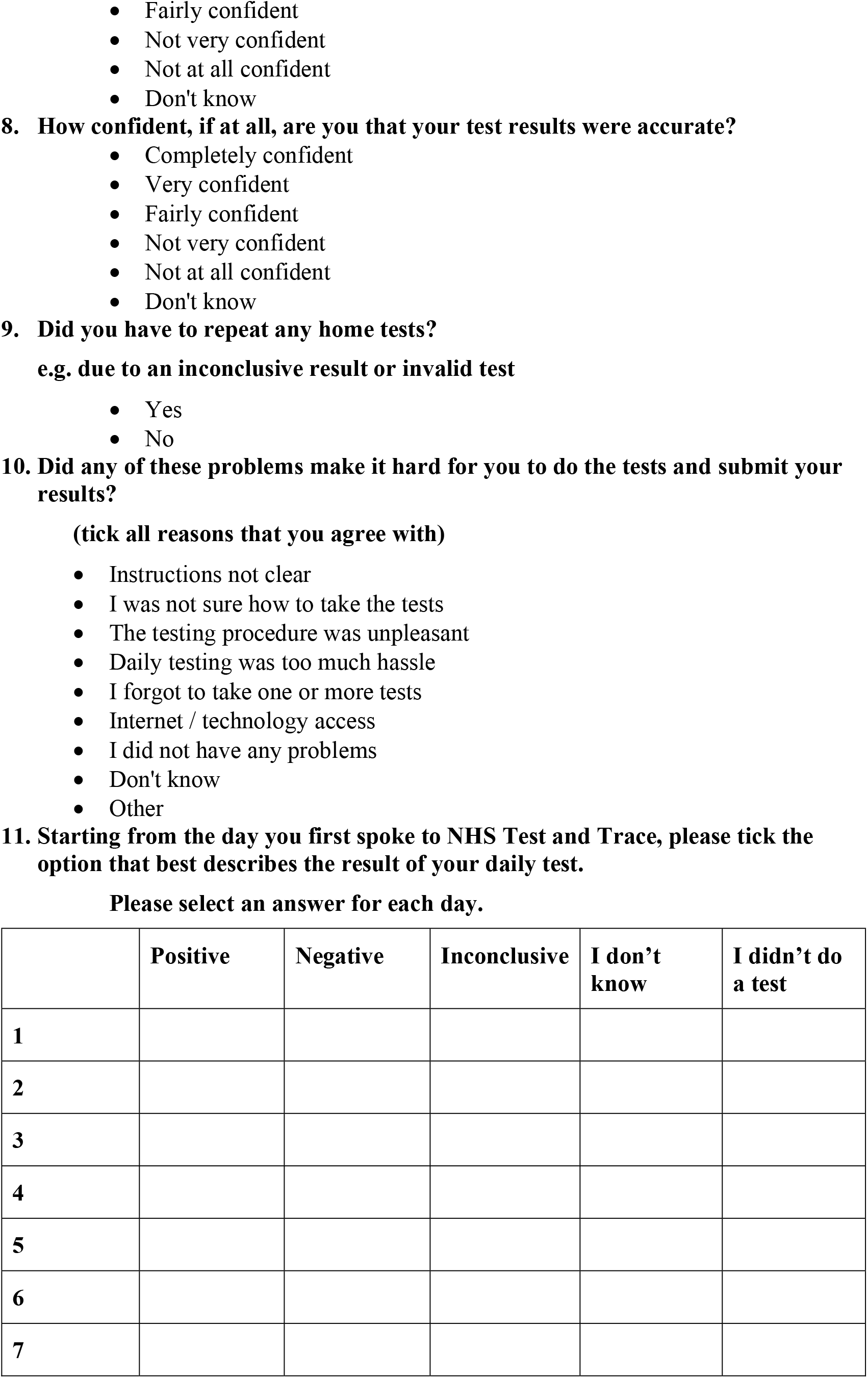
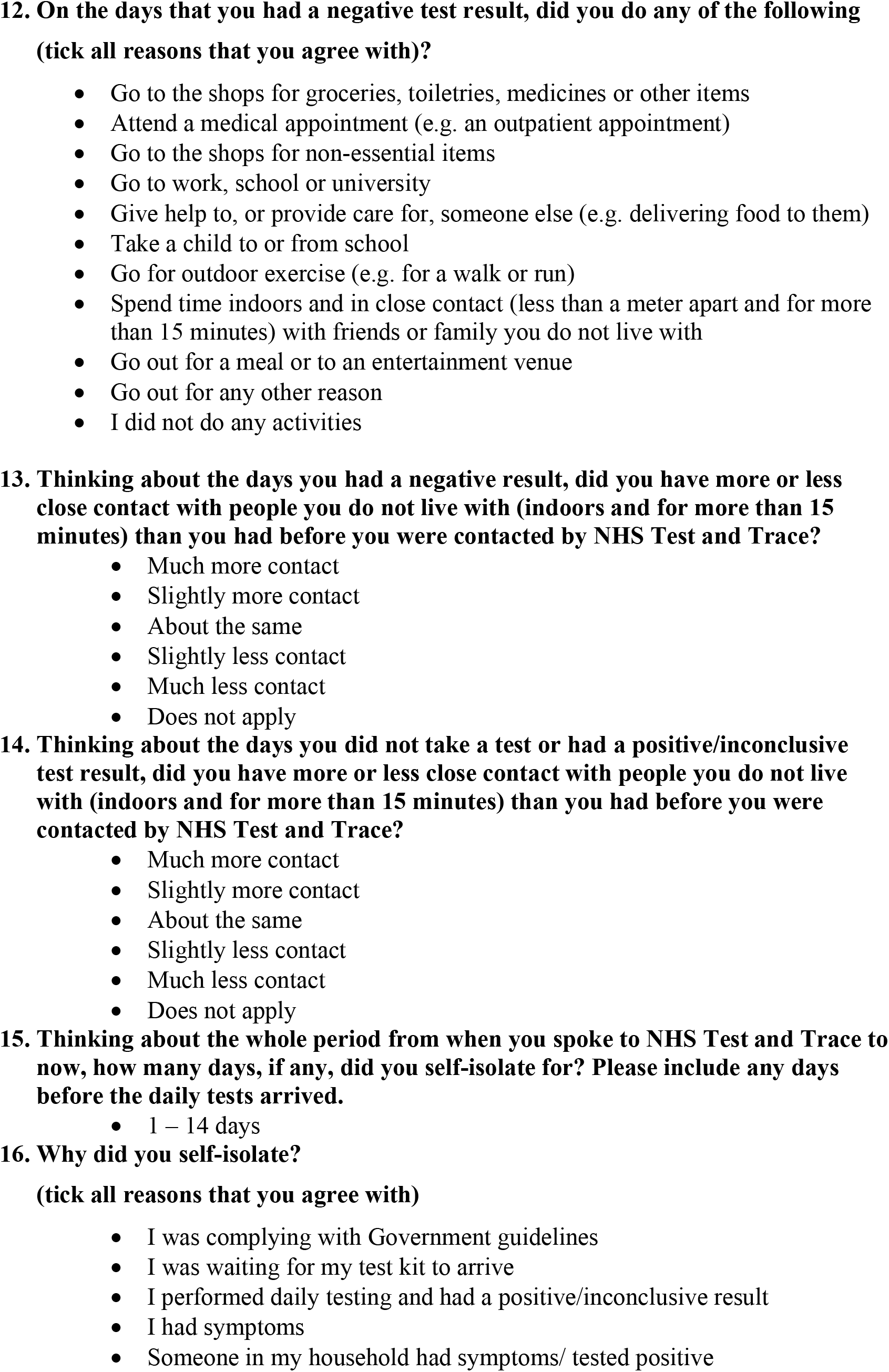
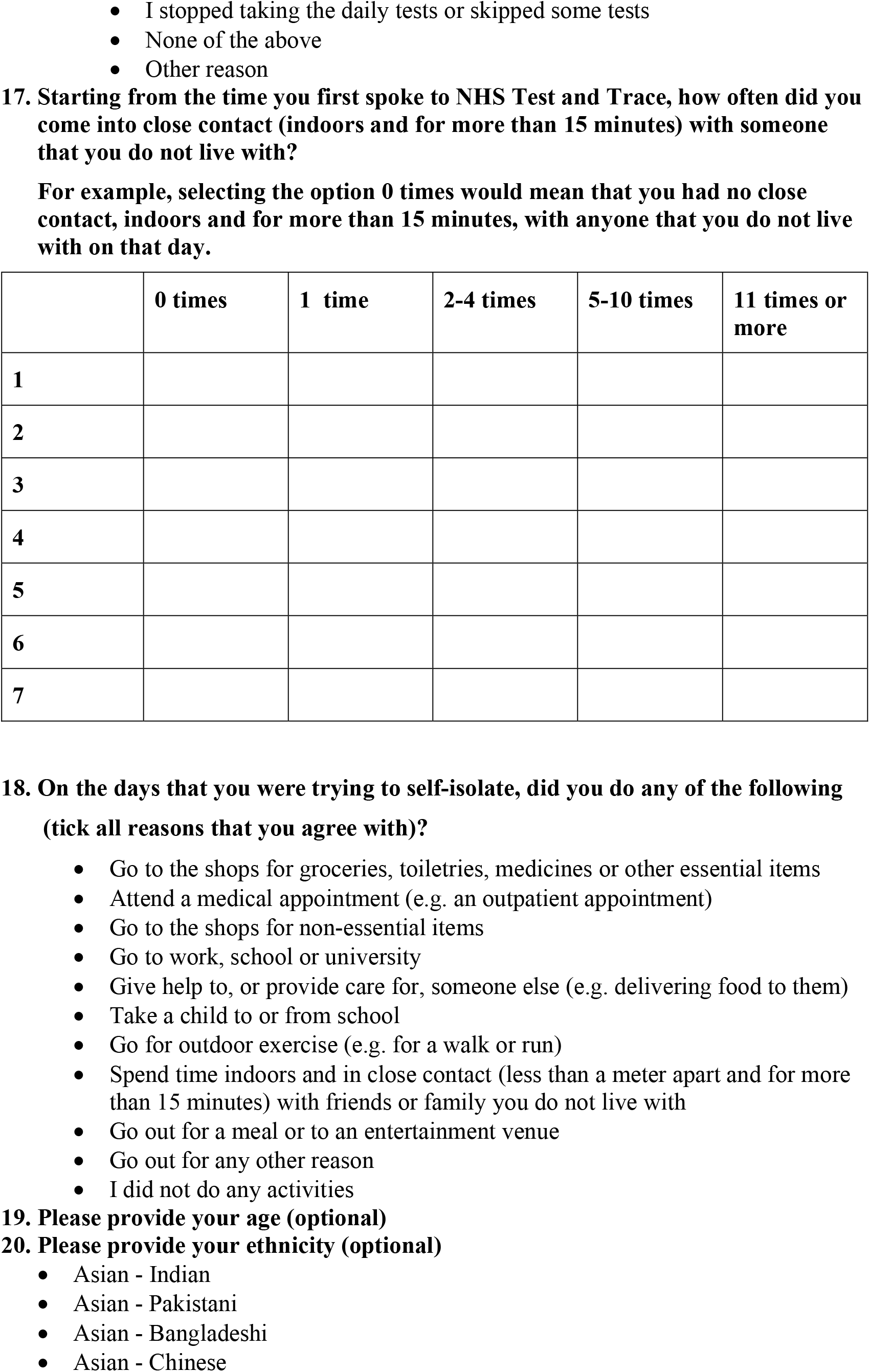
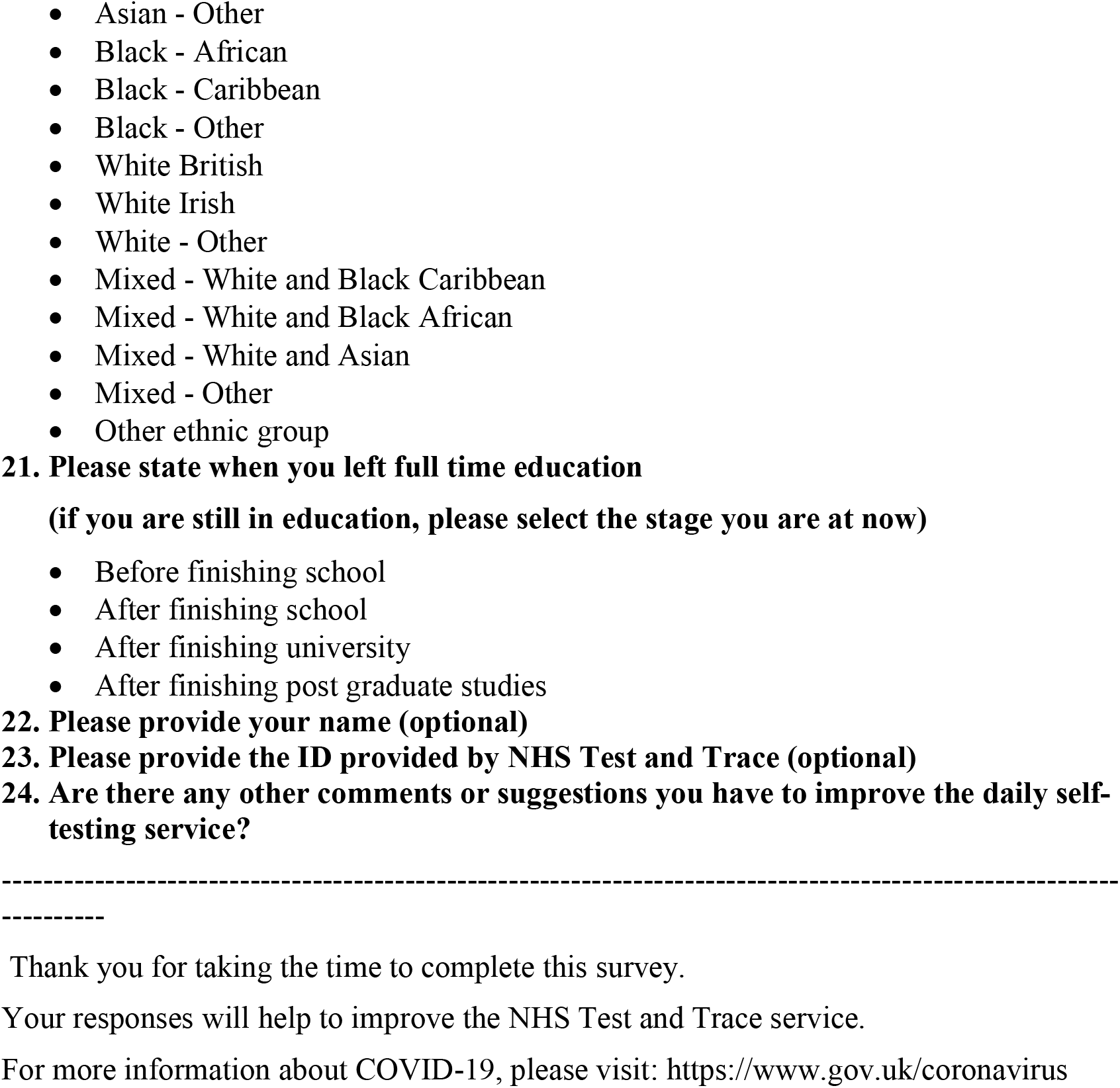
Survey questions and answers

**Supplementary table 2.**
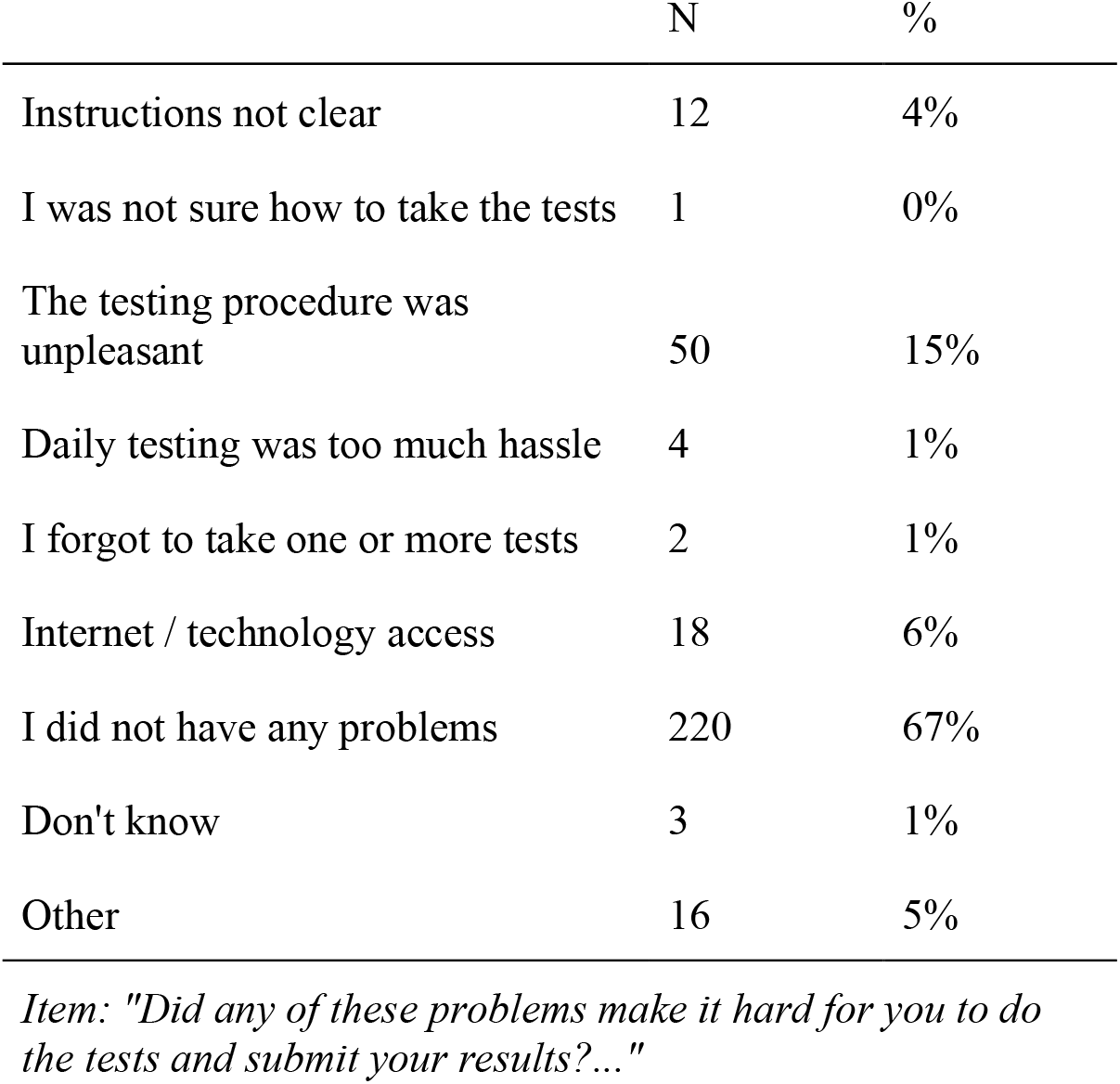
Issues with daily testing

|  | N | % |
| --- | --- | --- |
| Instructions not clear | 12 | 4% |
| I was not sure how to take the tests | 1 | 0% |
| The testing procedure was unpleasant | 50 | 15% |
| Daily testing was too much hassle | 4 | 1% |
| I forgot to take one or more tests | 2 | 1% |
| Internet / technology access | 18 | 6% |
| I did not have any problems | 220 | 67% |
| Don't know | 3 | 1% |
| Other | 16 | 5% |

*Item: "Did any of these problems make it hard for you to do the tests and submit your results?..."*

**Supplementary table 3.** Why did you decide to complete daily tests rather than self-isolate for 10 days?

| Item | Whole<br>sample | % | Ethnic<br>minority | % | White | % | Secondary<br>education | % | Higher<br>education | % |
| --- | --- | --- | --- | --- | --- | --- | --- | --- | --- | --- |
| I felt it was compulsory / I had no choice | 3 | 0% | 2 | 3% | 1 | 0% | 1 | 0% | 1 | 0% |
| It sounded quite easy to do | 175 | 17% | 13 | 16% | 151 | 17% | 54 | 16% | 97 | 17% |
| It would be difficult to self-isolate/ did not want to self-isolate | 81 | 8% | 5 | 6% | 72 | 8% | 36 | 10% | 41 | 7% |
| I needed to go out to work | 38 | 4% | 5 | 6% | 31 | 3% | 8 | 2% | 26 | 5% |
| My friends and family wanted me to do it | 16 | 2% | 2 | 3% | 14 | 2% | 7 | 2% | 6 | 1% |
| My employer wanted me to do it | 3 | 0% | 0 | 0% | 2 | 0% | 2 | 1% | 1 | 0% |
| I wanted to know if I had the virus | 226 | 22% | 15 | 19% | 195 | 22% | 80 | 23% | 122 | 22% |
| I wanted to meet up with friends/family | 21 | 2% | 0 | 0% | 21 | 2% | 7 | 2% | 11 | 2% |
| I know other people who were getting tested | 10 | 1% | 3 | 4% | 7 | 1% | 5 | 1% | 4 | 1% |
| To help beat the virus in my area | 206 | 20% | 11 | 14% | 180 | 20% | 66 | 19% | 117 | 21% |
| I was concerned that I might have coronavirus | 91 | 9% | 9 | 11% | 76 | 9% | 32 | 9% | 46 | 8% |
| I needed to know if I was infected so I could protect vulnerable people that I live with or meet regularly | 133 | 13% | 11 | 14% | 113 | 13% | 38 | 11% | 73 | 13% |
| None of these reasons | 2 | 0% | 0 | 0% | 1 | 0% | 0 | 0% | 2 | 0% |
| Other reason | 28 | 3% | 3 | 4% | 25 | 3% | 9 | 3% | 13 | 2% |

Note. This item was also asked elsewhere in the participant recruitment that was not part of this survey, so numbers reported elsewhere may differ

